# Married Women’s Decision-Making Autonomy on Maternal and Neonatal Healthcare Utilization and Associated Factors in Debretabor, Northwest Ethiopia

**DOI:** 10.1101/2020.08.31.20185579

**Authors:** Azmeraw Ambachew Kebede, Getachew Azeze Eriku, Birhan Tsegaw Taye, Wagaye Fentahun Chanie

## Abstract

**Background:** Women’s decision-making autonomy is very crucial for the improvement of maternal, neonatal, and child healthcare utilization. However, there is limited evidence on women’s decision-making autonomy on maternal and neonatal health in the study area. Therefore, this study assessed married women’s decision-making autonomy on maternal and neonatal healthcare utilization and associated factors in Debretabor, Northwest Ethiopia.

**Methods:** A community-based cross-sectional study was conducted from October 1^st^ to 30^th^, 2019. A multi-stage sampling technique was used to select 730 married women. A structured, pretested, and interviewer-administered questionnaire was employed. Data were entered into epi info 7 and analyzed by SPSS version 23. Multivariable logistic regression model was fitted to identify factors associated with women’s decision-making autonomy on maternal and neonatal healthcare utilization. Odds ratio with 95 % confidence interval was computed to determine the level of significance.

**Result:** Three-fourths (75.1%) of women had higher decision-making autonomy regarding their health, neonatal health, and other social and economic aspects. Besides, the proportion of at least four antenatal visits, delivery at a health facility, postnatal checkup, knowledge of neonatal danger signs, and appropriate health-seeking practices for sick newborns among autonomous women were 52.1%, 56.1%, 71.4%, 32%, and 80% respectively. Age greater than 35 years (AOR=2.08; 95%CI: 1.19, 3.62), monthly income of 5000 ETB and above (AOR=3.1; 95%CI: 1.36, 7.07), husband involvement (AOR=2.36; 95%CI: 1.55, 3.43) and knowledge of neonatal danger signs (AOR=2.11; 95%CI: 1.4, 3.2) were factors independently associated with women’s decision-making autonomy on maternal and neonatal healthcare utilization.

**Conclusion:** Our findings show that women’s decision-making autonomy on maternal and neonatal healthcare utilization was optimal. Increasing the household income level through different means, the promotion of husband’s involvement, and increasing women’s knowledge of maternal and neonatal danger signs will have a great role in the improvement of women’s decision-making autonomy.

## Introduction

Women’s autonomy is defined as the ability of women to act independently on their particular health, children’s health, freedom of movement, and control over finance without requesting permission from somebody (1). Empowering women is very essential for any social and economic development of a country (2). Hence, women’s autonomy undoubtedly contributes to many health advantages for both the mother and their children. However, there is a dearth of strong description of the concept and obtaining data on the individual, household, and community level that shows all opportunities of women empowerment (3). Maternal and neonatal health provision needs a multi-sector approach which in turn requires a strong decision-making autonomy of women to reverse back the barriers at the household level (4).

Although maternal mortality has dropped by 2.9% every year between 2000-2017, there are still an estimated 295,000 maternal deaths and 2.6 million neonatal deaths in the year 2017 (5,6). The strong decision-making power of women is vital to dropping this higher magnitude. Because, limited women’s decision-making autonomy impedes maternal healthcare utilization such as antenatal care (ANC), postnatal care (PNC), and delivery at a facility (7). Besides, lower autonomy of women affects the socio-economic, emotional, fertility decision, contraceptive use, and the sexual life of the women (8,9). Notably, decisions made at the household level affect not only the welfare of the individual but also the surrounding community even to the country level (10).

Studies show that the decision-making autonomy of women is low, specifically in developing countries. However, scaling up the women’s role in a decision leads to better uptake of healthcare access, poverty reduction, and household economic growth (1,10,11). In developing countries, women play an important role in the beneficence of the family, but they essentially have seen as an ordinary homemaker (12). Literature in Ethiopia shows that maternal and neonatal health coverage is low (13,14). Recent data revealed that 43%, 48%, and 34% of women had four and above ANC visits, give birth at a health facility and postnatal check respectively (15). According to the 2016 Ethiopian demographic health survey (EDHS 2016), only 11-18% of women were involved in making decisions alone and 66-68% together with their husband or partner (16). Also, the neonatal mortality rate and the maternal mortality ratio of Ethiopia are 30 per 1000 live births (15) and 412 per 100,000 live births respectively (16).

So far, studies in Nigeria and parts of Ethiopia including Bale zone, Ambo town, Southern Ethiopia and analysis of 2011 EDHS data revealed that women’s decision-making autonomy was low; which was 38.9% (11), 41.4% (1), 55.6% (17), 58.4% (14) and 54% (18) respectively. Besides, studies from Iran, Nigeria and elsewhere in Ethiopia including Ambo town, Southern Ethiopia and Bale zone found that older maternal age, exposure to mass media, higher socioeconomic status, higher educational status, higher family size and knowledge of maternal and child health were positively associated with women’s decision-making autonomy (1,8,11,14,17).

Improving maternal and neonatal health is one of the government concerns both nationally and globally that comprises the third component of sustainable development goal (SDG) (19). Since improving the optimal health of both the mother and her neonate and typically decreasing the maternal and neonatal mortality as low as possible by assessing factors affecting maternal and neonatal healthcare utilization, specifically on the autonomy of women will undoubtedly have a significant role in the achievement of the 3^rd^ SDG. Even though a few kinds of researches were conducted on the decision-making autonomy of women, some published studies failed to address some variables like husband involvement in maternal, neonatal, and child health (MNCH) services and knowledge of the maternal and neonatal illness. Moreover, most of the published studies tried to correlate the decision-making autonomy of women with maternal healthcare utilization only, leaving back what factors affect their decision-making autonomy.

In Ethiopia, there is a significant burden of maternal and neonatal death due to low healthcare utilization (15,16). Promoting the decision-making autonomy of women is the mainstay to increase maternal and neonatal healthcare utilization. Therefore, this study assessed married women’s decision-making autonomy on maternal and neonatal healthcare utilization and associated factors in Debretabor, Northwest Ethiopia.

## Methods

### Study design, setting and period

A community-based cross-sectional study was conducted from October 1^st^ to 30^th^, 2019 in Debretabor, Northwest Ethiopia. The town is located at 665 km Northwest of Addis Ababa (the capital city of Ethiopia) and 103 km Northeast of Bahir Dar (the capital city of Amhara regional state) and it is the capital city of South Gondar Zone. Currently, the town has a total population of 63,563, of whom 31,863 (54.8 %) are female. An estimated 19,327 are women of the reproductive-age group. About 4317 (6.8%) are under five. Moreover, the town has one general hospital, three health centers, 6 Health posts, and 6 private clinics serving the community (Debretabor administrative town report, unpublished data).

### Study population

All married women who had an infant age of one year and below and residing for at least six months in the selected ‘kebeles were included in the study. Women who were seriously ill or mentally ill throughout the data collection period were excluded.

### Sample size determination & sampling procedures

The sample size for this study was determined by using a single population proportion formula by considering the following assumptions:- the proportion of women’s decision-making autonomy on maternal and neonatal healthcare utilization 58.4% (14), 95% level of confidence and 5% margin of error.

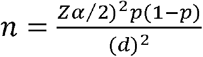

Where n= required sample sizes

α= level of significance
z= standard normal distribution curve value for 95% confidence level= 1.96
p= proportion of women’s decision-making autonomy on maternal and child healthcare utilization
d= margin of error
Therefore, 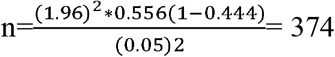

Finally, considering a 10% non-response rate and a design effect of 2 (since multi-stage sampling was employed). Accordingly, the final calculated sample size was 748. A multi-stage sampling technique was employed to select the study subjects. In the first stage, three ‘kebeles’ were selected by simple random sampling among the six ‘kebeles’, which is the smallest administrative unit in Ethiopia. The list of study participants was obtained from health extension workers and the sampling frame was designed by numbering the list of women using the registration book. Finally, a total of 748 participants were selected by a simple random sampling technique using a table of random generation. The calculated sample size was proportionally allocated to each ‘kebeles’.

### Measurement

**Women’s decision-making autonomy:** For this study, it was composited to higher decision-making autonomy (which was coded as ‘‘1’’) and lower decision-making autonomy (which was coded as ‘‘0’’) and three response variables (2= for women who were able to decide individually, 1=for women who were able to decide together with her husband and 0 = otherwise). Accordingly, women who scored above the mean were considered highly autonomous whereas women who scored below the mean were less autonomous (1,2). The women have questioned “who in your household decides (1) Healthcare for yourself (2), Healthcare for newborns and/ or children (3) Visit of family or relatives (4) To have additional children (5) Utilization of maternal and child healthcare services like ANC, PNC, and immunization (6) Large household purchases and consumptions (7) Intra-household resource allocation and purchases (8) Husband’s earning and (9) Cooking daily foods. The likely replies for each question were women alone, together with her husband or husband only.

### Operational definitions

**Decision-making autonomy:** Women who scored above the mean were considered highly autonomous whereas women who scored below the mean were less autonomous (1,2).

**Good knowledge of NDS:** Women who mentioned at least three neonatal danger signs among 12 neonatal danger signs (20).

**Poor knowledge NDS:** Women who mentioned less than three neonatal danger signs among 12 neonatal danger signs (20).

**Appropriate health-seeking practices:** Women who sought care for neonatal danger signs from well-qualified health professionals in governmental and/or private health facilities (21).

**Inappropriate health-seeking practices:** Women who sought care for neonatal danger sign other than qualified health professionals like purchasing over the counter drugs, treating by home remedies, seeking care from temples and traditional healers (21).

**Adequate ANC:** Women who had four or more ANC visits (22).

**Facility delivery:** Delivery in public or private hospitals and/ or clinics (22).

**Had PNC:** Women who received at least one postnatal checkup (22).

### Data collection tools and procedures

Data were collected using a structured interviewer-administered questionnaire. Three diploma data collectors and three supervisors, who have a bachelor of science in midwifery degree, trained about the interview technique collected and supervise the data.

### Data quality controls

The questionnaire was first prepared in English and translated to the local Amharic language and back to English to keep the consistency of the questionnaire. Before the actual data collection, pretest was done on 5% of the sample size at Adis Zemen town, Northwest Ethiopia to check the response, language clarity, understanding of data collectors and supervisors about the questionnaire, and appropriateness of the questionnaire. The required modification was done before the actual data collection. During the actual data collection period, the questionnaire was checked for completeness daily by the supervisors and the principal investigator.

### Data processing and analysis

Data were checked, coded, and entered into EPI INFO version 7, and were exported to SPSS version 23 for analysis. Descriptive statistics were used to present participants’ characteristics and decision-making autonomy of women. Binary logistic regression model was fitted to identify statistically significant explanatory variables and variables having a p-value of less or equal to 0.2 were included in the multivariable logistic regression model for controlling confounders. In multivariable logistic regression, a p-value of <0.05 with 95% CI for odds ratio was used to determine the significance association. Model fitness was checked using Hosmer and Lemeshow goodness of fittest.

### Ethics approval and consent to participate

The study was done following the Ethiopian Health Research Ethics Guideline and the declaration of Helsinki. Ethical clearance was obtained from the University of Gondar College of Medicine and Health Sciences ethical review committee. A formal letter of administrative approval was obtained from Debretabor town health office. Written informed consent was taken from each of the study participants after a clear explanation of the aim of the study.

## Result

### Socio-demographic characteristics of study participants

A total of 730 married women were included in this study. Eighteen women were excluded from the study due to their incomplete data, giving 98% response rate. The mean age of the participants was 30 years (SD ±5.86). Most of the study participants (97.9%) were Orthodox Christian by religion. Almost half (49.2%) of the participants had accomplished diploma and above education. Half (50.8%) of women were housewives by occupation. Regarding the occupation of the husband 406 (55.6%) were government employees and two-thirds (66.7%) of them had completed college and above education [**Table 1]**.

**Table 1:**
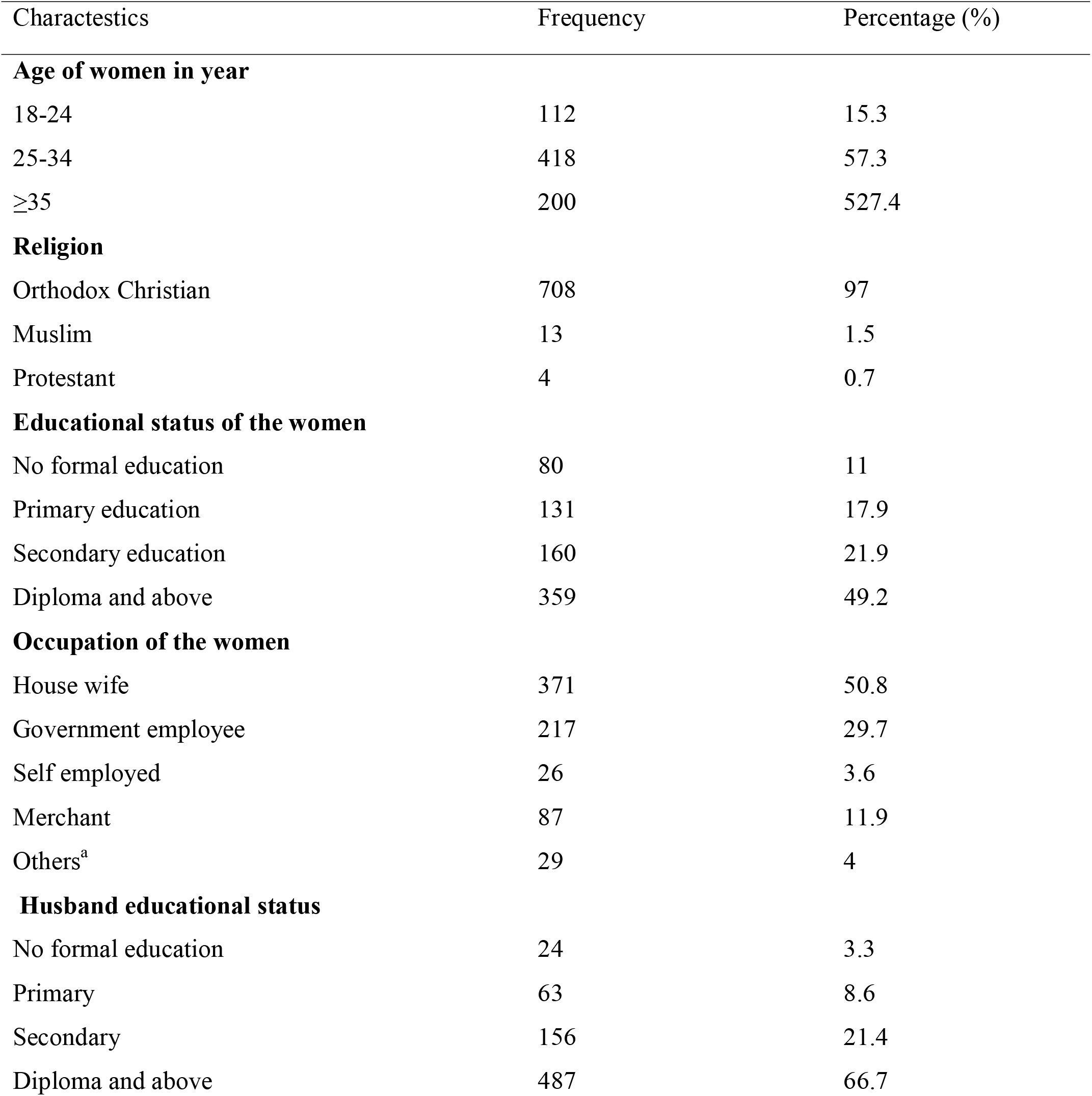
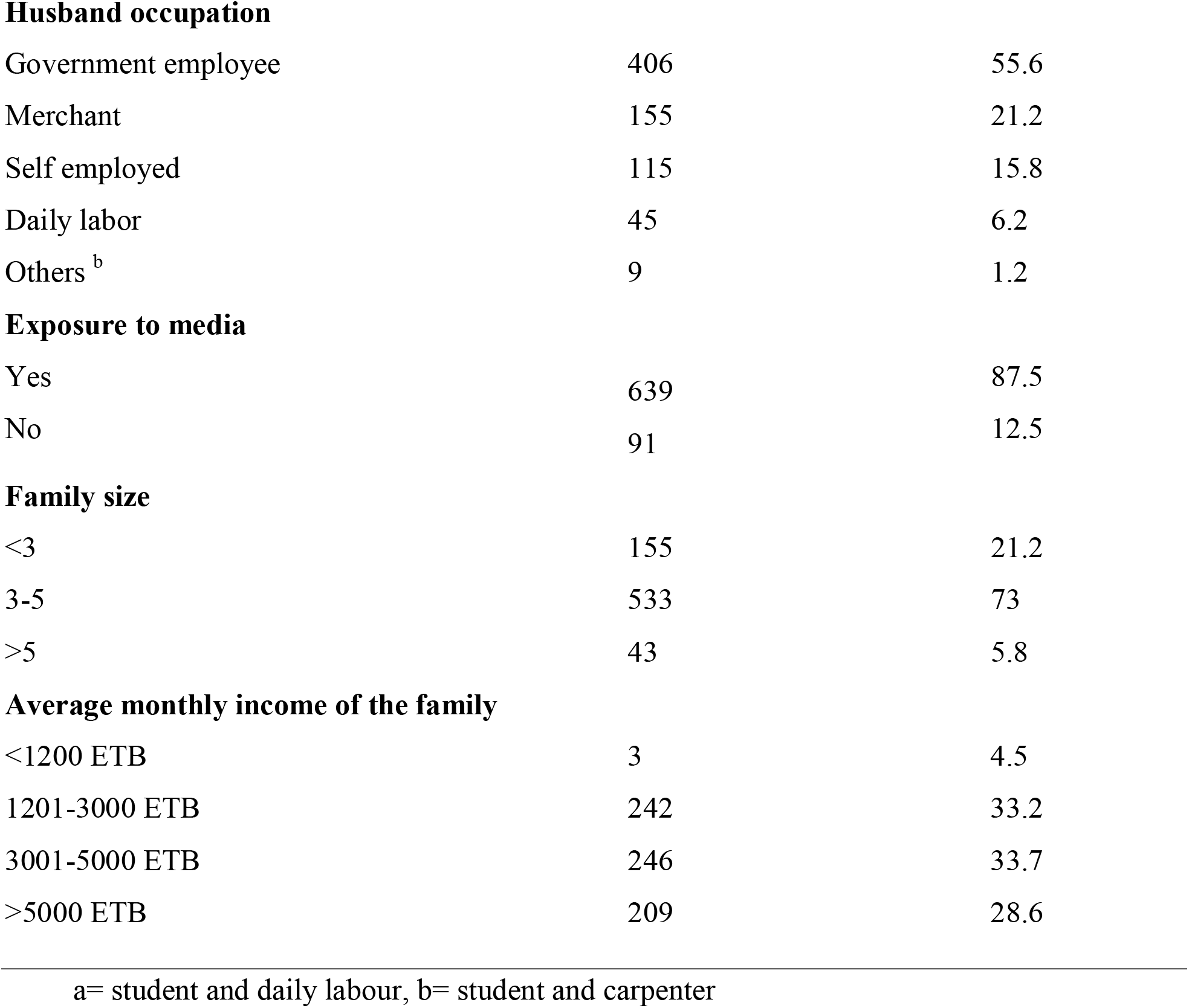
Socio-demographic characteristics of study participant in Debretabor, Northwest Ethiopia, 2019 (n=730)

| Characterstics | Frequency | Percentage (%) |
| --- | --- | --- |
| **Age of women in year** |  |  |
| 18-24 | 112 | 15.3 |
| 25-34 | 418 | 57.3 |
| ≥35 | 200 | 527.4 |
| **Religion** |  |  |
| Orthodox Christian | 708 | 97 |
| Muslim | 13 | 1.5 |
| Protestant | 4 | 0.7 |
| **Educational status of the women** |  |  |
| No formal education | 80 | 11 |
| Primary education | 131 | 17.9 |
| Secondary education | 160 | 21.9 |
| Diploma and above | 359 | 49.2 |
| **Occupation of the women** |  |  |
| House wife | 371 | 50.8 |
| Government employee | 217 | 29.7 |
| Self employed | 26 | 3.6 |
| Merchant | 87 | 11.9 |
| Others<sup>a</sup> | 29 | 4 |
| **Husband educational status** |  |  |
| No formal education | 24 | 3.3 |
| Primary | 63 | 8.6 |
| Secondary | 156 | 21.4 |
| Diploma and above | 487 | 66.7 |
| **Husband occupation** |  |  |
| Government employee | 406 | 55.6 |
| Merchant | 155 | 21.2 |
| Self employed | 115 | 15.8 |
| Daily labor | 45 | 6.2 |
| Others <sup>b</sup> | 9 | 1.2 |
| **Exposure to media** |  |  |
| Yes | 639 | 87.5 |
| No | 91 | 12.5 |
| **Family size** |  |  |
| <3 | 155 | 21.2 |
| 3-5 | 533 | 73 |
| >5 | 43 | 5.8 |
| **Average monthly income of the family** |  |  |
| <1200 ETB | 3 | 4.5 |
| 1201-3000 ETB | 242 | 33.2 |
| 3001-5000 ETB | 246 | 33.7 |
| >5000 ETB | 209 | 28.6 |

a= student and daily labour, b= student and carpenter

### Reproductive history and maternity healthcare service-related characteristics

From the total study participants, more than two-thirds (68.2%) of women had a parity of two to four. The majority of women (97.5%) had at least one ANC visit in their recent pregnancy, of whom, only 59.6% of women completed four ANC visits. Four-fifths (80.3%) of women gave birth at governmental health institutions for their recent delivery. Most (94%) of women had at least one postnatal visit for their most recent delivery [**Table 2]**.

**Table 2:**
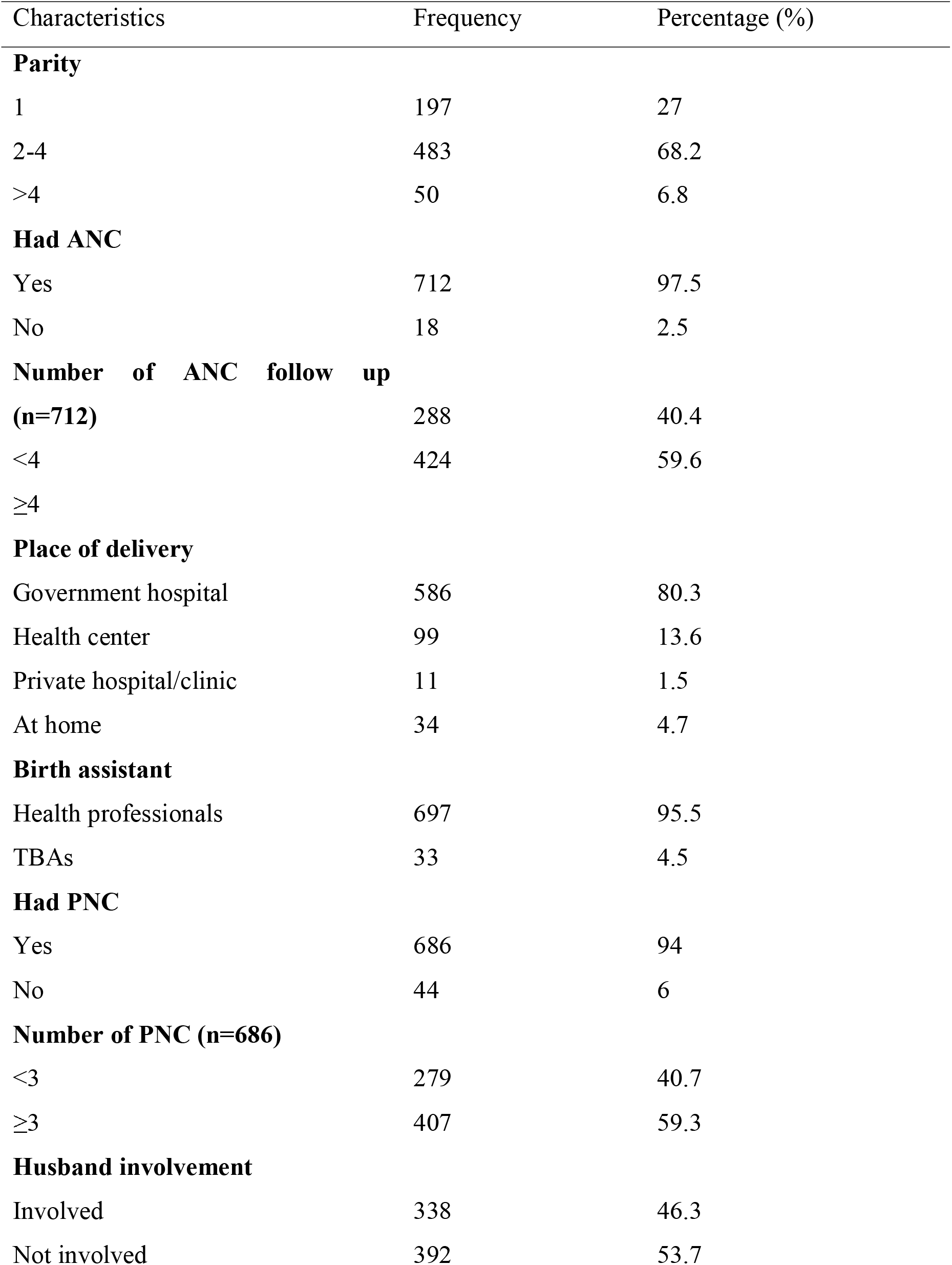
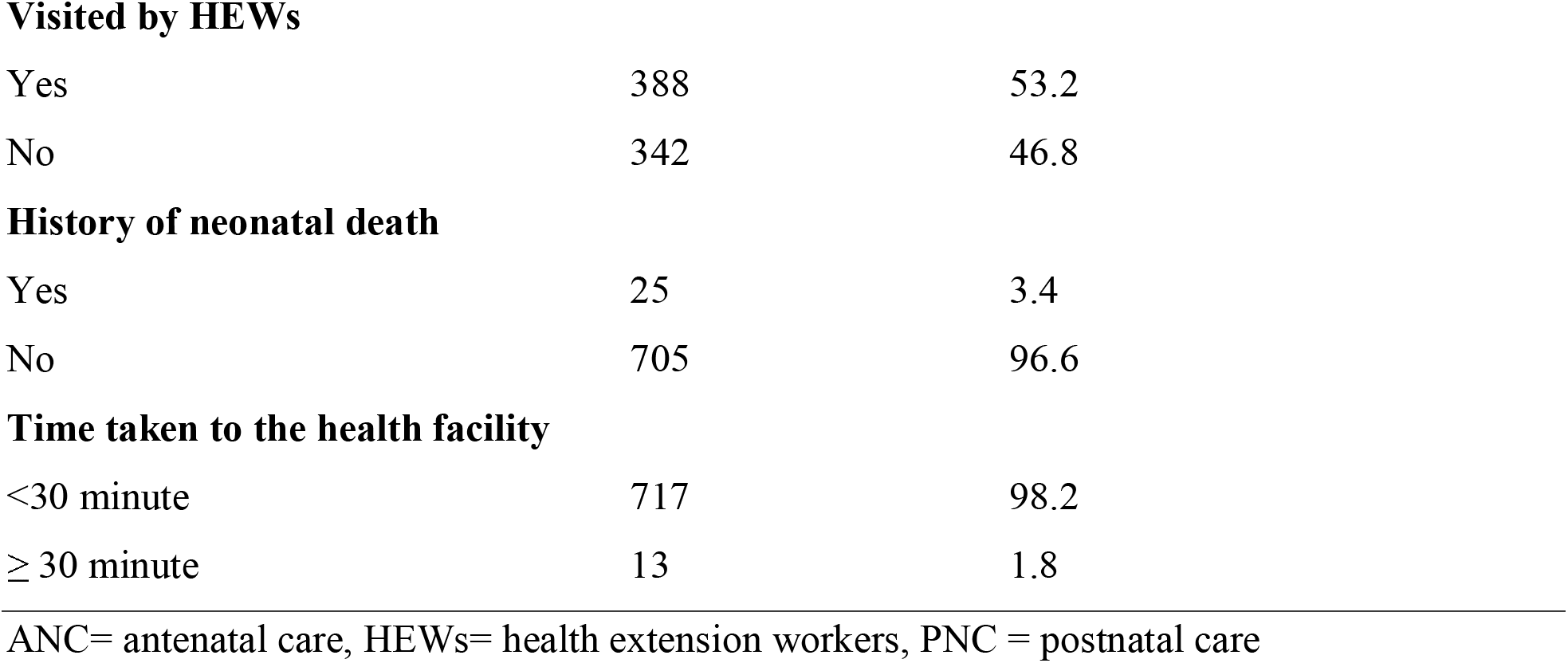
Reproductive history ad maternity healthcare service-related characteristics of study participant in Debretabor, Northwest Ethiopia, 2019 (n=730)

| Characteristics | Frequency | Percentage (%) |
| --- | --- | --- |
| **Parity** |  |  |
| 1 | 197 | 27 |
| 2-4 | 483 | 68.2 |
| >4 | 50 | 6.8 |
| **Had ANC** |  |  |
| Yes | 712 | 97.5 |
| No | 18 | 2.5 |
| **Number of ANC follow up (n=712)** | 288 | 40.4 |
| <4 | 424 | 59.6 |
| ≥4 |  |  |
| **Place of delivery** |  |  |
| Government hospital | 586 | 80.3 |
| Health center | 99 | 13.6 |
| Private hospital/clinic | 11 | 1.5 |
| At home | 34 | 4.7 |
| **Birth assistant** |  |  |
| Health professionals | 697 | 95.5 |
| TBAs | 33 | 4.5 |
| **Had PNC** |  |  |
| Yes | 686 | 94 |
| No | 44 | 6 |
| **Number of PNC (n=686)** |  |  |
| <3 | 279 | 40.7 |
| ≥3 | 407 | 59.3 |
| **Husband involvement** |  |  |
| Involved | 338 | 46.3 |
| Not involved | 392 | 53.7 |
| **Visited by HEWs** |  |  |
| Yes | 388 | 53.2 |
| No | 342 | 46.8 |
| **History of neonatal death** |  |  |
| Yes | 25 | 3.4 |
| No | 705 | 96.6 |
| **Time taken to the health facility** |  |  |
| <30 minute | 717 | 98.2 |
| ≥ 30 minute | 13 | 1.8 |

ANC= antenatal care, HEWs= health extension workers, PNC = postnatal care

### Women’s decision-making autonomy at the household level

The overall decision-making autonomy of women was found to be 75.1% (95%CI: 72.1, 78.1). About 72.1% of women had the joint decision to visit the health facilities for their health when they become sick. More than four-fifths (84.8%) of the study participants decide with their husbands to take sick newborns to the health facility. More than three-fourths (77.1%) and almost three-fourths (74.7%) of the participants had a joint decision with their husband for large household purchases and small intrahousehold resource allocation respectively [**Table 3]**.

**Table 3:**
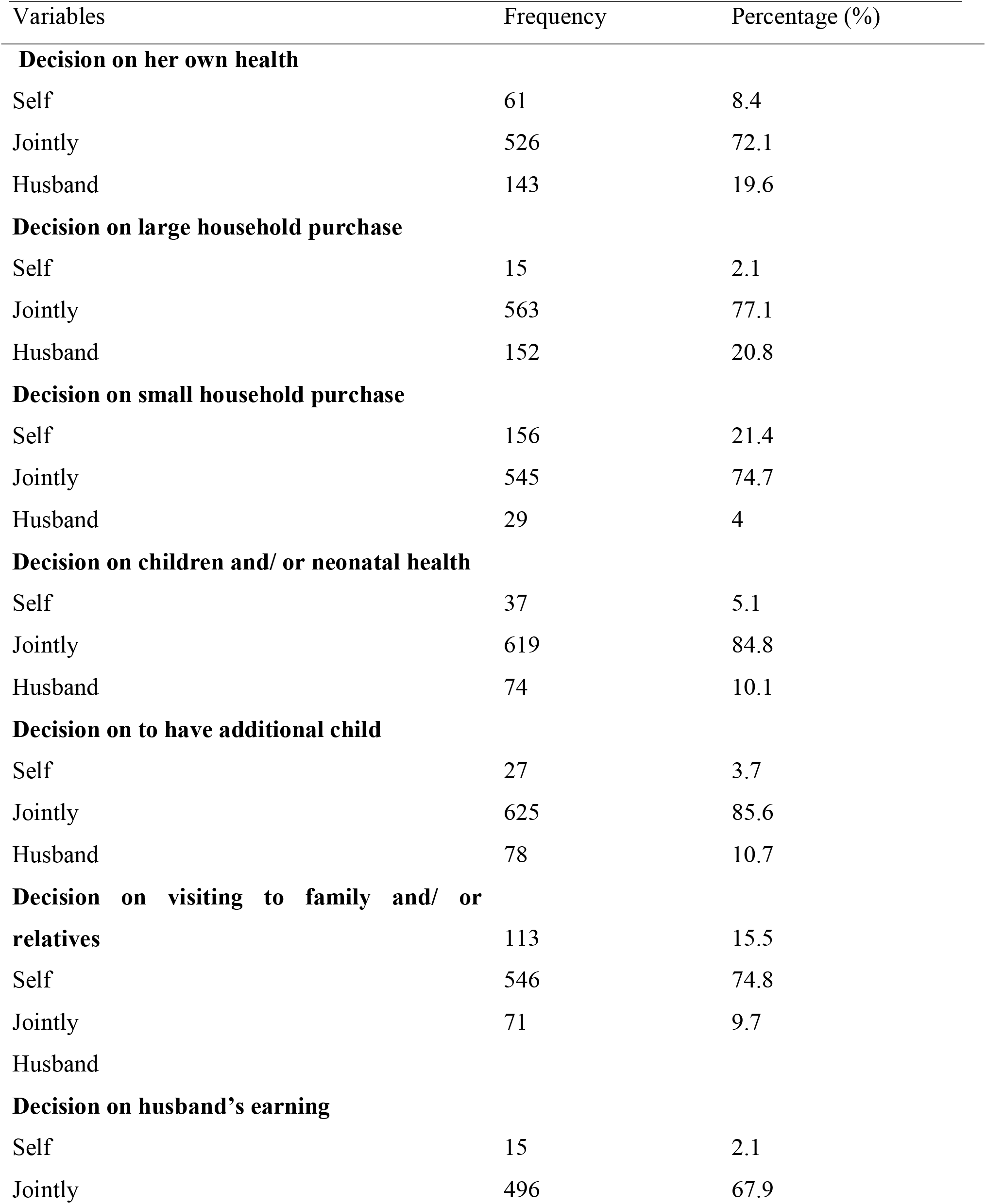
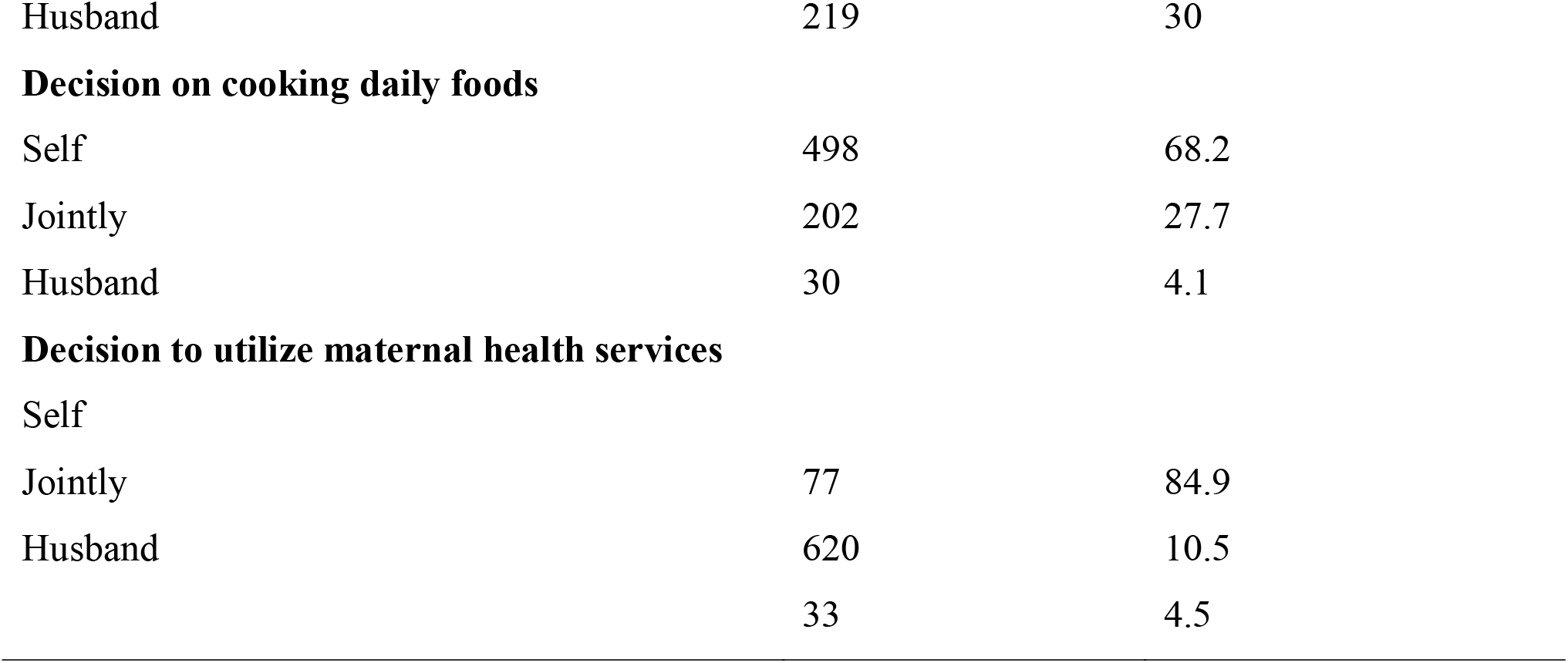
Women’s household decision-making autonomy characteristics, in Debretabor, northwest Ethiopia, 2019.

### Decision-making status of women and MNCH service utilization

The proportion of at least four ANC visits, delivery at a health facility, postnatal checkup, knowledge of neonatal danger signs, and appropriate health-seeking practices for sick neonates among autonomous women were 52.1%, 56.1%, 71.4%, 32%, and 80% respectively.

### Factors associated with women’s decision-making autonomy on maternal and neonatal healthcare utilization

From the multivariable logistic regression analysis age of the women, monthly income of the family, knowledge of NDS, and husband’s involvement in MNCH related activities had an association with women’s decision-making autonomy.

Those women greater than 35 years were two (AOR=2.08; 95%CI: 1.19, 3.62) times more likely to have had higher decision-making autonomy compared to their counterparts.

The odds of having higher decision-making autonomy among women who had a monthly income of 5000 ETB and above was three times higher compared to women who had a monthly income of less than 1200 ETB (AOR=3.10; 95% CI: 1.36, 7.07).

Similarly, those women who got husband involvement in MNCH related activities were 2.36 times (AOR=2.36; 95%CI: 1.60, 3.47) more likely to be autonomous compared to those women who didn’t have husband support.

This study also revealed that those women who had adequate knowledge of neonatal danger signs were two (AOR=2.11; 95%CI: 1.4, 3.2) times more likely to have had higher decision-making autonomy compared to women who had poor knowledge of newborn danger signs [**Table 4]**.

**Table 4:**
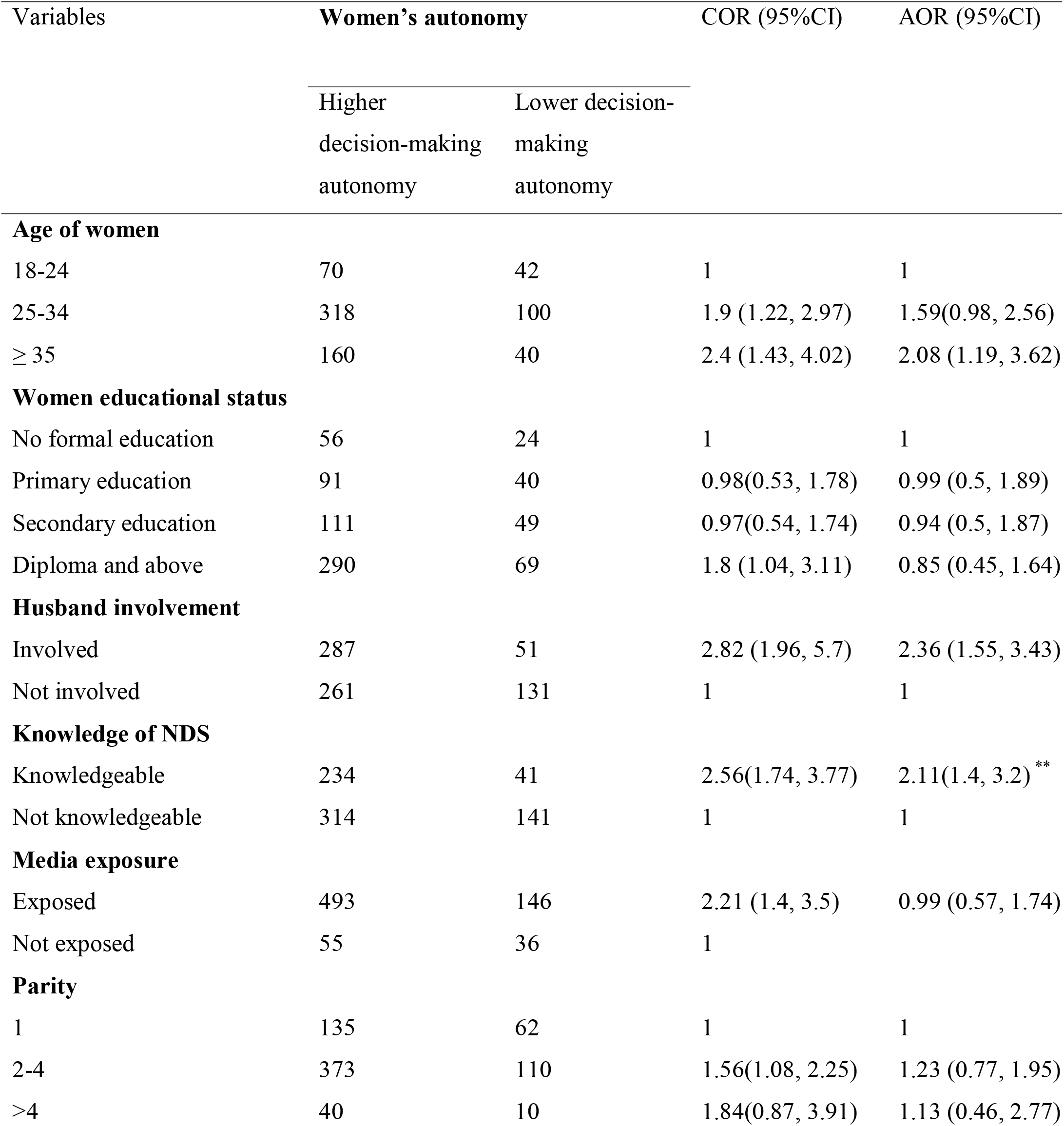
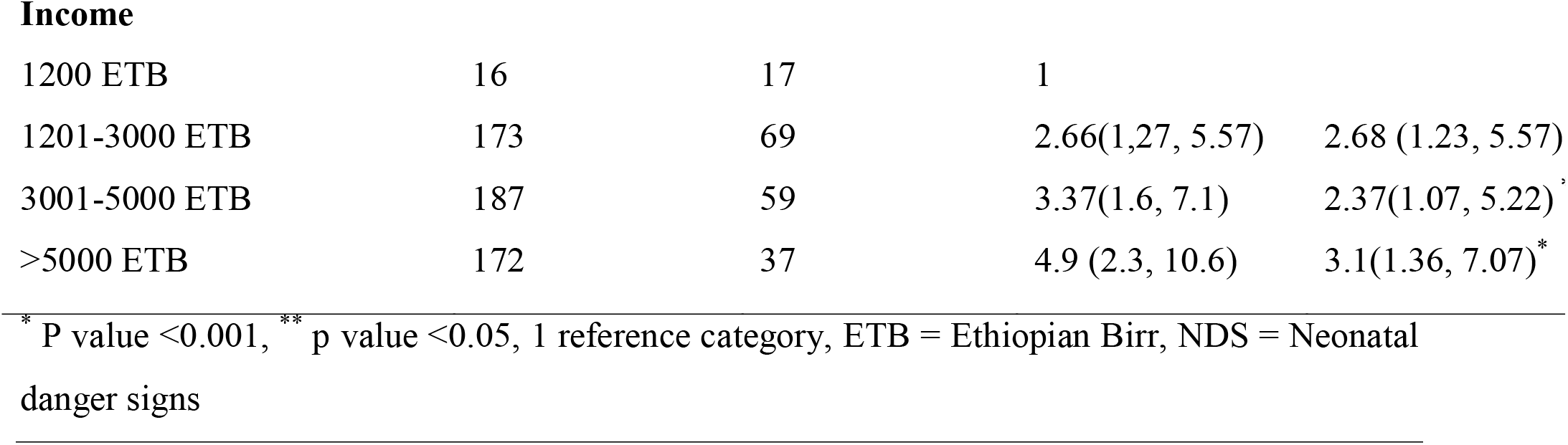
Bivariable and multivariable logistic regression analysis of factors associated with women’s decision-making autonomy among women who had an infant age of one year and below in Debretabor, Northwest Ethiopia, 2019 (n=730).

| Variables | Women's autonomy |  | COR (95%CI) | AOR (95%CI) |
| --- | --- | --- | --- | --- |
|  | Higher decision-making autonomy | Lower decision-making autonomy |  |  |
| **Age of women** |  |  |  |  |
| 18-24 | 70 | 42 | 1 | 1 |
| 25-34 | 318 | 100 | 1.9 (1.22, 2.97) | 1.59(0.98, 2.56) |
| ≥ 35 | 160 | 40 | 2.4 (1.43, 4.02) | 2.08 (1.19, 3.62) |
| **Women educational status** |  |  |  |  |
| No formal education | 56 | 24 | 1 | 1 |
| Primary education | 91 | 40 | 0.98(0.53, 1.78) | 0.99 (0.5, 1.89) |
| Secondary education | 111 | 49 | 0.97(0.54, 1.74) | 0.94 (0.5, 1.87) |
| Diploma and above | 290 | 69 | 1.8 (1.04, 3.11) | 0.85 (0.45, 1.64) |
| **Husband involvement** |  |  |  |  |
| Involved | 287 | 51 | 2.82 (1.96, 5.7) | 2.36 (1.55, 3.43) |
| Not involved | 261 | 131 | 1 | 1 |
| **Knowledge of NDS** |  |  |  |  |
| Knowledgeable | 234 | 41 | 2.56(1.74, 3.77) | 2.11(1.4, 3.2) ** |
| Not knowledgeable | 314 | 141 | 1 | 1 |
| **Media exposure** |  |  |  |  |
| Exposed | 493 | 146 | 2.21 (1.4, 3.5) | 0.99 (0.57, 1.74) |
| Not exposed | 55 | 36 | 1 |  |
| **Parity** |  |  |  |  |
| 1 | 135 | 62 | 1 | 1 |
| 2-4 | 373 | 110 | 1.56(1.08, 2.25) | 1.23 (0.77, 1.95) |
| >4 | 40 | 10 | 1.84(0.87, 3.91) | 1.13 (0.46, 2.77) |
| **Income** |  |  |  |  |
| 1200 ETB | 16 | 17 | 1 |  |
| 1201-3000 ETB | 173 | 69 | 2.66(1.27, 5.57) | 2.68 (1.23, 5.57) |
| 3001-5000 ETB | 187 | 59 | 3.37(1.6, 7.1) | 2.37(1.07, 5.22) <sup>‡</sup> |
| >5000 ETB | 172 | 37 | 4.9 (2.3, 10.6) | 3.1(1.36, 7.07) <sup>*</sup> |

<sup>\*</sup> P value <0.001, <sup>\*\*</sup> p value <0.05, 1 reference category, ETB = Ethiopian Birr, NDS = Neonatal danger signs

## Discussion

Women’s decision-making autonomy is very important for the wellbeing of the family, particularly for the improvement of maternal and neonatal health in resource-limited countries like Ethiopia. This study assessed married women’s decision-making autonomy on maternal and neonatal healthcare utilization among women who had an infant of one year and below in Debretabor, Northwest Ethiopia. Three-fourths of women had higher decision-making autonomy on maternal and neonatal healthcare utilizations. Furthermore, this study found that the proportion of at least four ANC visits, delivery at a health facility, at least one postnatal checkup, good knowledge of neonatal danger signs and appropriate health-seeking practice for sick newborns among autonomous women was 52.1%, 56.1%, 71.4%, 32%, and 80% respectively.

We found that the overall women’s decision-making autonomy on maternal and neonatal healthcare utilization was 75.1% (95%CI: 72.1, 78.1), which was higher than the previous studies conducted in Nigeria (11) and in parts of Ethiopia including Bale zone (1), Ambo town (17), Southern Ethiopia (14) and analysis from EDHS 2011 (18). This variation might be due to the difference in the time gap, cultural beliefs, and the study population’s socio-demographic characteristics. The present study was conducted from the urban population, in which the habit of attending information and realizing it is higher in urban areas. Besides, our study was conducted in a specific area, whereas the result from Nigeria was from the national demographic health survey.

The current study found that maternal age greater than 35 years were two times more likely to have had higher decision-making autonomy compared to their counterparts. A similar result was reported from Southern Ethiopia, in which women greater than 30 years were more likely to be participating in the decisions on their health (14). This might be due to, as age gets increased their educational status may have also increased and respect between couples will be increased with age.

Adequate knowledge of neonatal danger sign plays a great role in the decision making autonomy of women on maternal and neonatal health. Thus, this study revealed that women who had adequate knowledge of neonatal danger signs were two times more likely to have had higher decision-making autonomy compared to women who had poor knowledge of neonatal danger signs. This finding is consistent with a study conducted in Bale zone, Ethiopia, in which women who had adequate knowledge of maternal and child health were independently associated with women’s autonomy on maternal and child healthcare utilization (1). This may be because having good knowledge of maternal and neonatal health enforces women to challenge their husbands because they comprehend the seriousness of the illness.

Similarly, this study found that monthly income had a direct association with women’s decision-making autonomy on maternal and neonatal healthcare utilization. Women who had a monthly income of 5000 ETB and above were three times more likely to have had a higher decision-making autonomy for their health, neonatal health, and other social and economic activities compared to women who had a monthly income of less than 1200 ETB. This finding is in agreement with a study conducted in Southern, Ethiopia (14). This could be explained by economically capable women who are more likely to use communication devices (like television, radio, and magazine) and utilize maternal and neonatal health information than the poor population.

Moreover, women who got husbands involved in MNCH related activities were 2.36 times more likely to have had higher decision-making autonomy compared to women who hadn’t get husband involvement. Studies elsewhere reported that husband involvement encourages women’s participation in social and economic domains (23), associated with women’s use of skilled maternal and neonatal health services (24), better intra-spousal communication, birth preparedness and readiness for complications and utilization of maternal health services which in turn improve women’s decision-making autonomy and a shared decision between couples (11,25). The other explanation might be the certainty of the women will be increased because of being helped and encouraged by their husbands and accordingly increased shared decision.

## Conclusion

Women’s decision-making autonomy on maternal and neonatal healthcare utilization was optimal. Older maternal age, higher economic status, adequate knowledge of neonatal danger signs, and husband’s involvement in MNCH were the factors that contribute women to have had higher decision-making autonomy on maternal and neonatal healthcare utilization. There is a need for integrated work to increase women’s decision-making autonomy, thereby increasing the utilization of maternal and neonatal healthcare.

### Limitation of the study

Social desirability bias may not be eliminated. However, a better way of understanding was created by trained data collectors about the importance of the study and their participation can play an incredible effect on the benefits of the study findings.

## Data Availability

The datasset is available and any one can obtain based resanonable request

https://www.elaz.com

**Abbreviations**
AOR: Adjusted Odds Ratio
ANC: Antenatal Care
CI: Confidence Interval
COR: Crude Odds Ratio
EDHS: Ethiopian Demographic Health Survey
ETB: Ethiopian Birr
MNCH: Maternal, Neonatal, and Child Health
NDS: Neonatal Danger Signs
PNC: Postnatal Care
SDG: Sustainable Development Goal
SPSS: Statistical Package for Social Science

## Acknowledgments

We would like to thank the University of Gondar for providing study ethical clearance and financial support. Our thankfulness also goes to all data collectors and study participants. We are glad to Debretabor Town Health Office for giving us a permission letter.

## Author contributions

All the authors had significant involvement in the conception and designing the study, acquisition of data, analysis, and interpretation of data, took part in drafting the article, revising the article, gave final approval of the version to be published, have agreed on the journal to which the article has been submitted and agree to be accountable for all aspects of the work.

## Availability of data and material

The datasets collected and analyzed for the current study are available from the corresponding author and can be obtained on a reasonable request.

## Disclosure

The authors declare that they have no conflict of interest in this work.

## Notes

### Competing Interest Statement

The authors have declared no competing interest.

### Clinical Protocols

http://www.azmu.com

### Funding Statement

no exteranal funding

### Author Declarations

The study was conducted in accordnce with the delaration of Helsinki and ethical clearance was obtained from university of Gondar IRB

